# Inflammatory Burden, Structural Exposure, and Racial Disparities in Incident Heart Failure: Evidence from the Multi-Ethnic Study of Atherosclerosis

**DOI:** 10.64898/2026.09.28.26364238

**Authors:** Félix E. Rivera-Mariani, Tashoy Campbell, Emily Solomon

**Affiliations:** College of Arts and Sciences, Lynn University, Boca Raton, FL, USA; RIPLRT Institute, Fort Lauderdale, FL, USA; Department of Microbiology and Immunology, School of Medicine, University of Puerto Rico–Medical Sciences Campus, San Juan, PR, USA

**Keywords:** heart failure, inflammation, health equity, structural determinants, MESA, biomarkers

## Abstract

**Background:** Racial disparities in incident heart failure (HF) are well documented, yet the extent to which systemic inflammatory burden—shaped by structural determinants of environmental exposure—accounts for these disparities remains unquantified in multi-ethnic prospective cohorts.

**Methods:** Among 6785 participants in the Multi-Ethnic Study of Atherosclerosis (enrolled 2000–2002; 4 racial/ethnic groups) followed for approximately 16 years, we estimated associations of 4 prespecified baseline inflammatory and hemostatic biomarkers (interleukin-6 [IL-6], C-reactive protein [CRP], fibrinogen, and D-dimer; per 1-SD increment) with incident HF (485 events) using Cox proportional hazards models. A composite inflammatory burden index was derived by principal components analysis. Race × biomarker multiplicative interactions were tested with Holm correction. Sequential biomarker adjustment quantified the percentage of the Black–White log-hazard difference explained relative to a clinical-only model. Robustness to ambient air pollution (PM_2.5_, NO_2_) was assessed in a geocoded subcohort.

**Results:** All 4 biomarkers were independently associated with incident HF (IL-6: hazard ratio [HR], 1.20 [95% CI, 1.08–1.34]; D-dimer: 1.18 [1.06–1.30]; CRP: 1.14 [1.02–1.27]; fibrinogen: 1.13 [1.02–1.24]). The composite index (first principal component, 53.7% of variance explained) yielded an HR of 1.25 (1.12–1.39) per 1-SD increase. No multiplicative race × biomarker interaction was detected (all Holm-corrected P=1.0), indicating that the per-SD inflammatory hazard was directionally consistent across groups. Sequential biomarker adjustment reduced the Black–White log-hazard difference by 57.5%, a pattern consistent with—but not sufficient to prove—attenuation by inflammatory pathways. Associations were robust to PM_2.5_/NO_2_ co-adjustment.

**Conclusions:** Inflammatory and hemostatic burden partially explains the Black–White disparity in incident HF. The null multiplicative interaction pattern indicates that per-unit biological risk is equitable across groups, consistent with differential structural exposure—rather than differential susceptibility—as a primary driver of disparity.

**Clinical Perspective:** **What Is New?**

- Baseline interleukin-6, C-reactive protein, fibrinogen, and D-dimer were each independently associated with incident heart failure in the Multi-Ethnic Study of Atherosclerosis, and a principal-components composite of the 4 markers outperformed any single marker.
- The per-SD hazard associated with inflammatory and hemostatic burden was consistent across the 4 racial and ethnic groups, whereas baseline burden was systematically higher among Black and Hispanic participants.
- Sequential adjustment for the 4-marker panel reduced the Black–White log-hazard difference for incident heart failure by 57.5%.

**What Are the Clinical Implications?**

- Inflammatory and hemostatic biomarkers may help identify individuals at elevated risk of heart failure who could benefit from intensified preventive strategies, although the incremental gain in discrimination beyond clinical risk factors was modest.
- Because per-unit biological risk was shared across groups while exposure burden differed, efforts to reduce racial disparities in heart failure should target the structural determinants that produce elevated inflammatory burden rather than presume group-specific biological susceptibility.
- Formal causal mediation and additive-interaction analyses are needed before inflammatory burden can be regarded as an established mediator of heart failure disparities.

## Introduction

Heart failure (HF) remains a leading cause of cardiovascular morbidity and mortality in the United States, affecting more than 6 million adults, with projections indicating a 46% increase in prevalence by 2030.^1^ Despite therapeutic advances, outcomes are marked by profound racial disparities driven by structural exposures rather than biological essentialism. In the Multi-Ethnic Study of Atherosclerosis (MESA), which followed 6,785 participants for about 16 years, the inequitable distribution of HF was stark: 153 events occurred among 1,880 Black participants (8.14%), compared with 200 events among 2,613 White participants (7.65%).

Hispanic participants experienced 99 events among 1,491 enrolled participants (6.64%), while Chinese American participants experienced 33 events among 801 enrolled participants (4.12%). These data reflect broader national trends in which Black individuals face significantly higher HF incidence and mortality rates than White counterparts, a gap rooted in the cumulative impact of residential segregation and unequal access to social determinants of health.^2,3^

Chronic systemic inflammation serves as a critical biological conduit through which structural inequities translate into adverse cardiac phenotypes. Interleukin-6 (IL-6) has been implicated in cardiac injury pathways in experimental models, where it promotes myocyte apoptosis and activates JAK-STAT3-mediated fibrosis; whether this sequence operates in humans with incident HF remains to be established.^4^ CRP further exacerbates myocardial injury by inducing endothelial dysfunction and complement-mediated myocardial depression.^5^ Simultaneously, the hemostatic system interacts with inflammatory cascades; fibrinogen serves as both an acute-phase reactant and a precursor to fibrin, promoting microvascular thrombosis and impairing diastolic filling.^6^ Finally, D-dimer, a terminal product of coagulation activation, signals systemic crosstalk between thrombosis and inflammation that is particularly relevant to the pathophysiology of heart failure with preserved ejection fraction.^7^ Together, these biomarkers represent a multisystemic inflammatory burden that may mediate racial disparities produced by structural exposures.

Structural racism—the systemic organization of mutually reinforcing institutions across housing, employment, credit, and environmental policy that collectively produces and sustains racial health inequity^8^—is the primary upstream determinant of differential inflammatory burden in MESA. Through residential segregation, Black and Hispanic communities are systematically concentrated in environments with elevated PM_2.5_ and NO_2_ exposure—a policy-driven geography of harm, not a product of biological predisposition.^9^ This environmental injustice, compounded by the persistent psychosocial stress of navigating systemic inequity, does not produce transient inflammation; it may recalibrate immune tone—the durable, structurally set inflammatory baseline. The current analysis operationalizes this construct as a single-timepoint composite biomarker, which serves as a proxy for, but does not directly measure, longitudinal inflammatory patterning. Rivera-Mariani et al. established a mechanistic precedent in Puerto Rico, demonstrating that environmental stressors, such as fungal spores and aeroallergens, induce sustained immune dysregulation.^10,11^ Within the MESA cohort, this manifests as a significantly higher baseline inflammatory burden in Black participants compared with White participants across all biomarkers studied, reflecting the cumulative physiological toll of structurally imposed environmental hazards.^9,12^

Prior landmark investigations, such as the Framingham Offspring Study, were conducted in predominantly White cohorts and did not report tests for race–biomarker interactions, limiting generalizability to diverse populations and precluding assessment of whether inflammatory burden varies across racial/ethnic groups. While these studies established associations, they did not quantify the proportion of racial disparities in HF incidence that is statistically explained by inflammatory burden, a necessary step toward understanding whether inflammation constitutes a mediating pathway. Furthermore, previous multi-ethnic analyses often lacked the geocoding rigor required to account for environmental confounding. MESA uniquely addresses these limitations through its multi-ethnic architecture and a high-accuracy geocode retention rate of 91.3%, enabling unprecedented co-adjustment for geocoded ambient air pollution and structural determinants.^13^

We tested three pre-specified hypotheses: (1) that each of four inflammatory and hemostatic biomarkers would independently predict incident HF; (2) that a composite inflammatory burden index would provide incremental discrimination (C-index and Brier score) beyond a clinical covariate model; (3a) that inflammatory burden would partially explain the Black–White HF disparity (sequential mediation); and (3b) that no multiplicative race × biomarker interaction would be detected, consistent with shared per-unit biological susceptibility under differential structural exposure.

## Methods

### Study Design and Population

MESA is a prospective longitudinal cohort study of 6,814 community-dwelling adults aged 45–84 years, free of clinically manifest cardiovascular disease at enrollment (2000–2002), recruited from six U.S. field sites across four self-identified racial/ethnic groups: White, Chinese American, Black, and Hispanic.^13^ The present analysis includes 6,785 participants with adjudicated follow-up (main analytic cohort) and 6,476 participants in the geocoded air pollution sub-cohort. Data were accessed under an executed Data Use Agreement (MESA public-use dataset v2025a). An exempt Lynn University IRB approval (Lynn IRB 23.26) was submitted to NHLBI-BioLINCC to access the data and be granted the Data Use Agreement.

### Outcome Ascertainment

Incident heart failure was centrally adjudicated by the MESA morbidity and mortality committee using standardized criteria that incorporated symptoms, signs, imaging, and physician diagnosis.^14, 15^ Time-to-event was measured in days from Exam 1 to the first adjudicated HF event, death, loss to follow-up, or administrative censoring.

### Exposure Variables

Four baseline (Exam 1) biomarkers—IL-6 (pg/mL), CRP (mg/L), D-dimer (μg/mL FEU), and fibrinogen (mg/dL)—were pre-specified as the primary confirmatory family prior to data analysis. IL-6, CRP, and D-dimer were log-transformed and standardized (z-scored), and fibrinogen was standardized on the untransformed scale. A composite inflammatory burden index was derived as the first principal component (PC1) of the four standardized biomarkers (variance explained: 53.7%; n = 6,567 complete), which was retained and z-scored. PCA was chosen to capture shared variance across biomarkers without imposing a pre-specified weighting structure, allowing the data to reveal the dominant inflammatory dimension. The variance explained by PC1 exceeded 50%, a commonly used threshold for retaining a single component, and was consistent with the hypothesis that inflammation and hemostasis constitute a unified systemic burden.

### Covariates

Fully adjusted models included age, sex, self-identified race/ethnicity, body mass index, systolic blood pressure, hypertension, diabetes mellitus, estimated glomerular filtration rate, smoking status and pack-years, education, income, baseline left ventricular ejection fraction, and MESA field site, all measured at Exam 1. BMI, hypertension, diabetes, and eGFR are plausibly downstream of chronic inflammatory burden; conditioning on these variables may attenuate biomarker associations (collider bias). Sensitivity analyses using minimally adjusted models (age, sex, and race only) were conducted to assess the magnitude of this bias (results in Table S5).

### Statistical Analysis

All statistical analyses were performed using R version 4.6.0 (R Foundation for Statistical Computing, Vienna, Austria). Cox proportional hazards models estimated hazard ratios (HRs) per 1-SD biomarker increment; the proportional hazards assumption was assessed using Schoenfeld residuals. Race × biomarker multiplicative interactions were tested with Wald tests of cross-product terms, with Holm’s method applied across the four-biomarker family.

Multiplicative interaction assesses whether the per-unit relative hazard varies by race, which is mechanistically informative. Additive interaction (RERI) assesses whether absolute risk differences vary by race, which is more directly relevant to equity science; RERI analyses are ongoing and will be reported in a companion manuscript. Benjamini–Hochberg q-values were reported in the discovery context. Missing biomarker data were addressed with multiple imputation (m = 20; mice package), and pooled estimates were compared with a complete-case analysis. To operationalize hypothesis (3), we used sequential biomarker adjustment to quantify the proportion of the Black–White log-hazard difference ‘explained’ by each biomarker, recognizing that this approach approximates but does not replace formal counterfactual mediation. Incremental discrimination was evaluated using Harrell’s C-index, time-dependent AUC, and the integrated Brier score across nested models. Environmental robustness was assessed by co-adjusting for PM_2.5_ and NO_2_ and by restricting geocode accuracy. A two-sided α = 0.05 governed all confirmatory claims. Dr Rivera-Mariani had full access to all the data in the study and takes responsibility for its integrity and the data analysis.

## Results

### Cohort Characteristics

Among 6,785 participants (38.5% White, 27.7% Black, 22.0% Hispanic, 11.8% Chinese American), 485 incident HF events occurred over approximately 16 years of follow-up. Event rates were highest among Black participants (8.14%), followed by White (7.65%), Hispanic (6.64%), and Chinese American (4.12%). Baseline inflammatory burden was systematically higher among Black and Hispanic participants across all four biomarkers: for example, the median standardized CRP was +0.22 among Black participants versus −0.11 among White participants (Table 1; Figure S1).

**Table 1.**
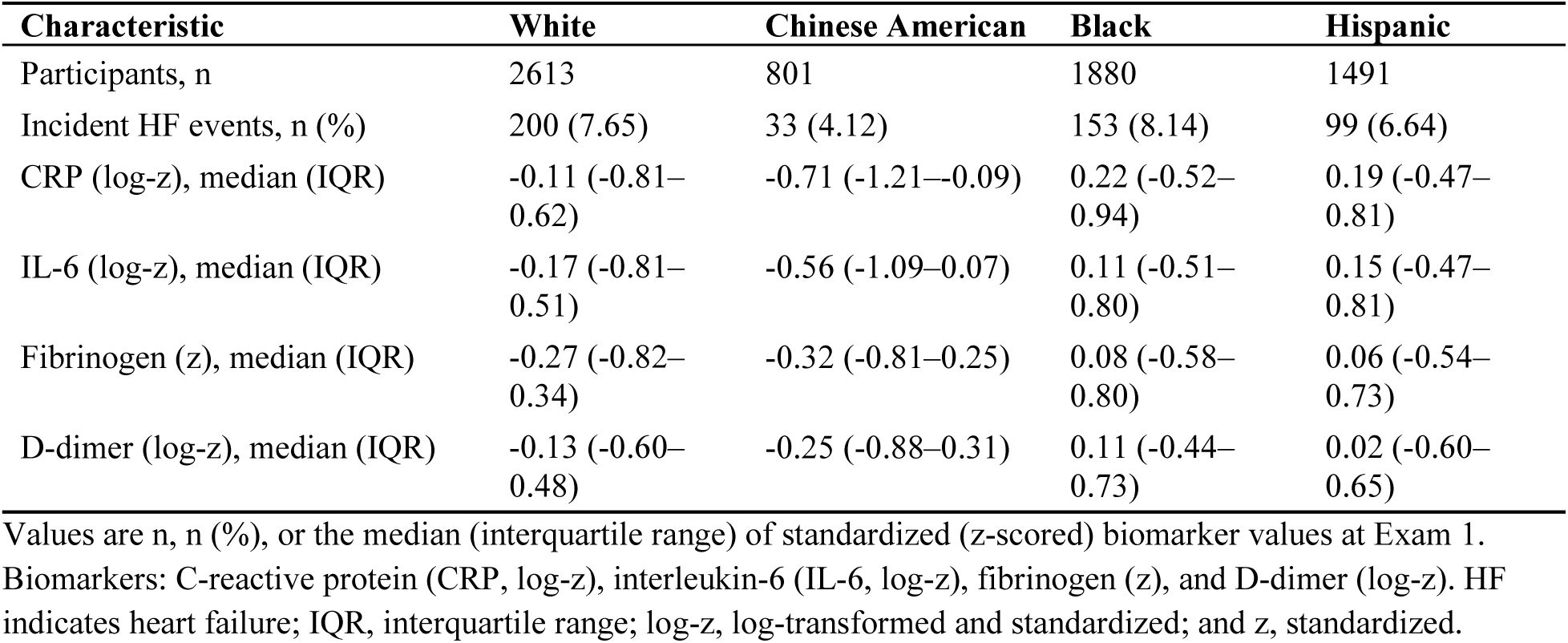
Baseline Characteristics and Standardized Biomarker Distributions of MESA Participants by Race/Ethnicity.

### Primary Biomarker Associations with Incident Heart Failure

All four prespecified biomarkers were independently associated with incident HF in fully adjusted models (Table 2; Figure 1A): IL-6, HR 1.20 (95% CI 1.08–1.34; P = 0.0010); D-dimer, HR 1.18 (1.06–1.30; P = 0.0017); CRP, HR 1.14 (1.02–1.27; P = 0.017); and fibrinogen, HR 1.13 (1.02–1.24; P = 0.015). These associations are modest in magnitude but consistent across markers, aligning with expectations for community-based biomarker studies. Multiple imputation yielded materially identical estimates (IL-6, 1.19; D-dimer, 1.18; CRP, 1.15; fibrinogen, 1.11), indicating robustness to missing-data mechanisms (Table S1).

**Figure 1.**
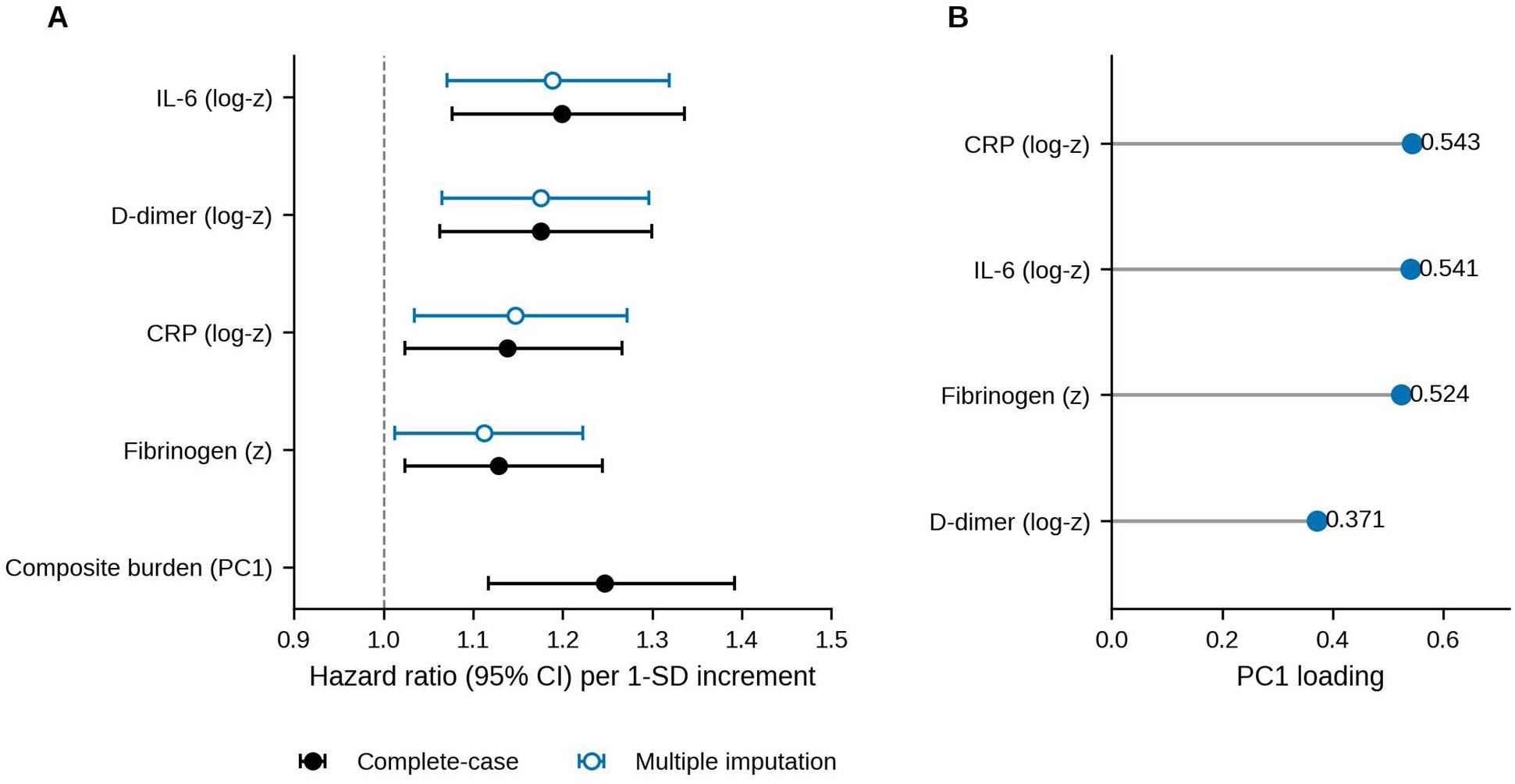
Adjusted hazard ratios for incident heart failure per 1-SD biomarker increment and loadings of the composite inflammatory burden index. **A**, Hazard ratios (95% CI) per 1-SD biomarker increment from fully adjusted Cox proportional hazards models (age, sex, race/ethnicity, body mass index, systolic blood pressure, hypertension, diabetes, estimated glomerular filtration rate, smoking, education, income, baseline left ventricular ejection fraction, and field site). Filled circles denote complete-case estimates (n=6178–6309; 442–449 events), and open circles denote multiple-imputation pooled estimates (m=20 imputed data sets; Rubin’s rules). The composite inflammatory burden index (first principal component [PC1]) is shown for comparison (n=6157; 440 events). The dashed vertical line indicates a hazard ratio of 1.0. **B**, PC1 loadings for the 4 Exam 1 biomarkers (variance explained, 53.7%; n=6567 participants with complete biomarker data). Approximately equal loadings for CRP, IL-6, and fibrinogen, with a moderate D-dimer loading, indicate a shared inflammatory dimension with a semi-independent hemostatic signal. Figures were generated in Python (matplotlib) from precomputed model summaries. CRP indicates C-reactive protein; IL-6, interleukin-6; log-z, log-transformed and standardized (z-scored); and z, standardized.

**Table 2.** Associations of Baseline Inflammatory and Hemostatic Biomarkers With Incident Heart Failure: Complete-Case and Multiple-Imputation Estimates.

| Biomarker | Participants,<br>n | Events,<br>n | Complete-case<br>HR (95% CI) | P value | Multiple-imputation HR<br>(95% CI) | P value |
| --- | --- | --- | --- | --- | --- | --- |
| IL-6 (log-z) | 6178 | 442 | 1.20 (1.08–1.34) | <0.001 | 1.19 (1.07–1.32) | 0.0012 |
| D-dimer (log-z) | 6309 | 449 | 1.18 (1.06–1.30) | 0.0017 | 1.18 (1.07–1.30) | 0.0013 |
| CRP (log-z) | 6304 | 448 | 1.14 (1.02–1.27) | 0.017 | 1.15 (1.03–1.27) | 0.0094 |
| Fibrinogen (z) | 6306 | 449 | 1.13 (1.02–1.24) | 0.015 | 1.11 (1.01–1.22) | 0.027 |
| Composite<br>inflammatory burden<br>index (PC1)* | 6157 | 440 | 1.25 (1.12–1.39) | <0.001 | — | — |

Hazard ratios (HRs) per 1-SD biomarker increment from fully adjusted Cox proportional hazards models (age, sex, race/ethnicity, body mass index, systolic blood pressure, hypertension, diabetes, estimated glomerular filtration rate, smoking, education, income, baseline left ventricular ejection fraction, and field site). Multiple imputation: m=20 data sets, Rubin's rules. P values from Wald tests. CI indicates confidence interval; CRP, C-reactive protein; IL-6, interleukin-6; log-z, log-transformed and standardized; PC1, first principal component; and z, standardized.
\*Composite index derived as the first principal component of the 4 standardized biomarkers (variance explained, 53.7%); complete-case analysis only.

### Composite Inflammatory Burden Index

PC1 of the four biomarkers explained 53.7% of the shared variance, with approximately equal loadings for CRP (0.543), IL-6 (0.541), and fibrinogen (0.524), and a moderate loading for D-dimer (0.371) (Figure 1B). The composite inflammatory burden index yielded a hazard ratio of 1.25 per 1-SD increase, modestly exceeding that of any individual biomarker (Table 2; Figure 1B). This supports its use as a summary measure of shared inflammatory/hemostatic burden, though the incremental gain in discrimination was small.

### Race × Biomarker Interaction

No multiplicative race × biomarker interaction was detected for any biomarker (global interaction P: IL-6, 0.346; D-dimer, 0.441; fibrinogen, 0.869; CRP, 0.987; all Holm-corrected P = 1.0; all BH q ≥ 0.88) (Table 3). Group-specific hazard ratios were directionally consistent across all four racial/ethnic groups (Figure S2), indicating that the per-SD inflammatory hazard is shared across groups.

**Table 3.** Race/Ethnicity × Biomarker Multiplicative Interaction Tests and Group-Specific Hazard Ratios.

| Biomarker | Global interaction P value | Holm-corrected P value | BH q value | Race/ethnicity | HR (95% CI) per 1-SD | P value | P value vs White |
| --- | --- | --- | --- | --- | --- | --- | --- |
| IL-6 (log-z) | 0.346 | 1.00 | 0.881 | White | 1.20 (1.03–1.41) | 0.020 | Reference |
|  |  |  |  | Chinese American | 1.02 (0.69–1.50) | 0.93 | 0.43 |
|  |  |  |  | Black | 1.11 (0.92–1.35) | 0.27 | 0.52 |
|  |  |  |  | Hispanic | 1.40 (1.12–1.74) | 0.0026 | 0.26 |
| D-dimer (log-z) | 0.441 | 1.00 | 0.881 | White | 1.28 (1.10–1.48) | 0.0014 | Reference |
|  |  |  |  | Chinese American | 1.25 (0.90–1.73) | 0.18 | 0.89 |
|  |  |  |  | Black | 1.10 (0.92–1.31) | 0.29 | 0.20 |
|  |  |  |  | Hispanic | 1.07 (0.87–1.32) | 0.53 | 0.17 |
| Fibrinogen (z) | 0.869 | 1.00 | 0.987 | White | 1.12 (0.96–1.31) | 0.13 | Reference |
|  |  |  |  | Chinese American | 1.26 (0.83–1.91) | 0.28 | 0.62 |
|  |  |  |  | Black | 1.08 (0.92–1.27) | 0.33 | 0.73 |
|  |  |  |  | Hispanic | 1.17 (0.98–1.41) | 0.088 | 0.72 |
| CRP (log-z) | 0.987 | 1.00 | 0.987 | White | 1.16 (0.99–1.36) | 0.063 | Reference |
|  |  |  |  | Chinese American | 1.14 (0.76–1.71) | 0.53 | 0.93 |
|  |  |  |  | Black | 1.12 (0.93–1.33) | 0.23 | 0.72 |
|  |  |  |  | Hispanic | 1.13 (0.90–1.41) | 0.30 | 0.83 |

### Attenuation of the Black–White Heart Failure Gap

In the clinical-only model, the Black–White HR was 0.91 (95% CI, 0.68–1.21). Sequential biomarker adjustment progressively attenuated the Black–White log-hazard difference: CRP explained 19.0% of excess risk; IL-6, 13.7%; fibrinogen, 25.4%; D-dimer, 50.3%; and the full panel, 57.5% (Table 4; Figure 2). D-dimer was the dominant individual mediator (50.3%), exceeding any single inflammatory marker. However, the full panel explained 57.5%, which is less than the arithmetic sum of the four individual estimates (58.1%), indicating that the biomarkers share overlapping mediation pathways. The full-panel estimate represents the net inflammatory burden–mediated effect after accounting for this overlap.

**Figure 2.**
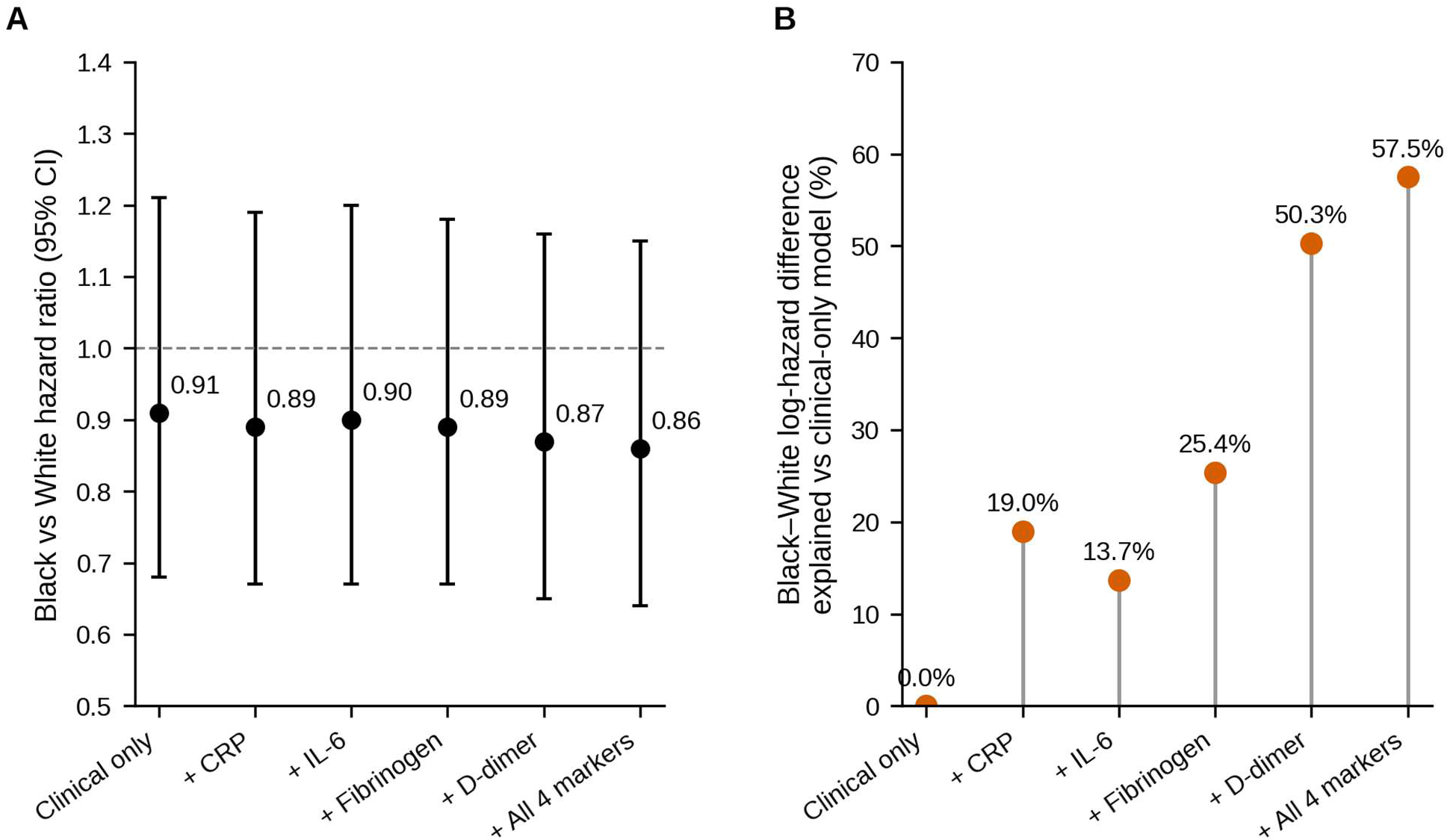
Sequential attenuation of the Black–White hazard ratio for incident heart failure with inflammatory biomarker adjustment. **A**, Black versus White hazard ratios (95% CI; Wald tests) from Cox models with each Exam 1 biomarker added sequentially to the clinical covariate set (clinical-only model: n=4151, 319 events; full 4-marker panel: n=4030, 310 events; per-model n in Table 4). The dashed line indicates a hazard ratio of 1.0. **B**, Percentage of the Black–White log-hazard difference explained relative to the clinical-only model (full panel, 57.5%; D-dimer alone, 50.3%). Abbreviations as in Figure 1.

**Table 4.** Sequential Attenuation of the Black–White Hazard Ratio for Incident Heart Failure by Inflammatory Biomarker Adjustment.

| Model | Participants, n | Events, n | Black vs White HR (95% CI) | P value | Excess risk explained, % |
| --- | --- | --- | --- | --- | --- |
| Clinical only | 4151 | 319 | 0.91 (0.68–1.21) | 0.52 | 0 (reference) |
| Clinical + CRP (log-z) | 4127 | 317 | 0.89 (0.67–1.19) | 0.44 | 19.0 |
| Clinical + IL-6 (log-z) | 4043 | 312 | 0.90 (0.67–1.20) | 0.47 | 13.7 |
| Clinical + Fibrinogen (z) | 4130 | 318 | 0.89 (0.67–1.18) | 0.42 | 25.4 |
| Clinical + D-dimer (log-z) | 4130 | 318 | 0.87 (0.65–1.16) | 0.33 | 50.3 |
| Clinical + all 4 biomarkers | 4030 | 310 | 0.86 (0.64–1.15) | 0.32 | 57.5 |

### Incremental Predictive Performance

Adding IL-6 and D-dimer to the clinical model increased the C-index from 0.785 to 0.790 and reduced the integrated Brier score from 0.037 to 0.036 (Table S2), consistent with the modest incremental discrimination typical of biomarkers in community cohorts.

### Environmental Robustness

Co-adjustment for PM_2.5_ and NO_2_ in the geocoded sub-cohort (n = 6,073) did not materially attenuate any biomarker association (IL-6, HR 1.18 [1.06–1.32]; D-dimer, 1.14 [1.03–1.27]; CRP, 1.14 [1.02–1.27]; fibrinogen, 1.14 [1.03–1.26]). Estimates were similarly stable under the geocode-accuracy restriction (n = 5,562) (Table S3).

## Discussion

In this prospective multi-ethnic cohort, all four pre-specified inflammatory and hemostatic biomarkers—IL-6, D-dimer, CRP, and fibrinogen—were independently associated with incident heart failure, with hazard ratios of 1.20, 1.18, 1.14, and 1.13 per 1-SD increase, respectively. To our knowledge, this is the first study to derive a principal-components composite of IL-6, CRP, fibrinogen, and D-dimer for HF prediction in a multi-ethnic cohort. Wang et al.^16^ constructed a multi-biomarker risk score for incident CVD but did not include D-dimer, did not use PCA, and did not test race–biomarker interactions. This work extends that framework by integrating hemostatic markers and evaluating whether inflammatory burden mediates racial disparities. These estimates align with prior evidence that serum IL-6 and CRP are elevated in persons who develop congestive heart failure.^17^ The composite inflammatory burden index provided a biologically coherent synthesis of the four markers, yielding a hazard ratio of 1.25 per 1-standard deviation increase, which exceeds that of any individual biomarker and extends the previously established multi-biomarker framework for cardiovascular risk prediction.^16^ The first principal component (PC1) accounted for 53.7% of the shared variance across the four markers. Its association with incident heart failure (HF) supports the conceptualization of HF risk as the cumulative manifestation of systemic immune dysregulation, or inflammatory immune tone, rather than the result of any single cytokine cascade. This baseline state of immune activation provides the context upon which structural exposures exert their effects.

Mechanistically, the biomarker panel encompasses two interconnected physiological systems. The classical inflammatory markers, interleukin-6 (IL-6) and C-reactive protein (CRP), represent the upstream cytokine signal. IL-6 induces hepatic JAK-STAT3-mediated acute-phase synthesis of CRP and fibrinogen, placing IL-6 at the apex of the inflammatory cascade.

Subsequently, the coagulation system is activated, as indicated by fibrinogen-to-fibrin conversion and elevated D-dimer levels. This process contributes to cardiac microvascular dysfunction and the heart failure with preserved ejection fraction phenotype.^18^ Within the composite index, the lower loading of D-dimer (0.371) compared with the inflammation trio (0.52–0.54) suggests a semi-independent hemostatic signal. This finding supports the view that the coagulation pathway represents a partially distinct mechanistic route to cardiac injury, separate from the inflammatory response.

The absence of a multiplicative race × biomarker interaction (global interaction P: IL-6, 0.346; D-dimer, 0.441; fibrinogen, 0.869; CRP, 0.987; all Holm-corrected P = 1.0) suggests shared biological susceptibility across groups. However, with 485 events distributed across four racial and ethnic groups, the study may have limited statistical power to detect moderate interactions, such as a hazard ratio (HR) of 1.2 versus 1.0. Approximate post-hc calculations suggest modest power for interaction detection; under an observed-sample approximation, power for a global race x biomarker interaction corresponding to a White HR of 1.0 versus Black HR of 1.3 was about 55%, indicating limited sensitivity to moderate heterogeneity. Therefore, the finding should be interpreted as the absence of a strong interaction rather than definitive evidence of homogeneity. This interpretation is consistent with the sufficient-cause framework, which distinguishes between identifying component causes that influence disease risk and designating any single factor as sufficient for disease.^19–21^

Multiplicative and additive interaction frameworks address different scientific questions. The relative excess risk due to interaction (RERI), also termed the interaction contrast ratio, assesses departure from additivity using risk ratios rather than absolute risks.^22^ RERI analysis is required by the equity science aims of this work because the additive scale addresses which groups bear more absolute disease burden—the public health question—rather than relative biological effect alone. Additive interaction analyses are ongoing and will be reported in a planned companion analysis.

The sequential mediation findings constitute a necessary first step in the equity science framework. By quantifying the proportion of the Black–White HF disparity explained by inflammatory burden (57.5%), these results establish that inflammation is a plausible mediator, though formal causal decomposition (planned via CMAverse) is required to satisfy counterfactual assumptions. Relative to the clinical-only model (Black vs. White HR 0.91), sequential biomarker adjustment progressively attenuated the Black–White log-hazard difference: CRP accounted for 19.0% of the excess risk; IL-6, 13.7%; fibrinogen, 25.4%; D-dimer, 50.3%; and the full inflammatory panel, 57.5%. D-dimer alone explained more of the mediated excess risk than the three classical inflammatory markers combined—a dominance that underscores the role of downstream coagulation activation in translating structural exposure into cardiac risk. These findings support conceptualizing inflammatory burden as a biological transducer of structural racism—consistent with higher baseline biomarker levels among Black and Hispanic participants and with shared per-unit risk across groups—rather than a biological property of race itself. They are also consistent with the weathering hypothesis, in which allostatic load differences between Black and White Americans widen across the life course under cumulative social stressors.^23^ Structural racism has been formally identified by the American Heart Association as a fundamental barrier to achieving cardiovascular health for all.^24^

Co-adjustment for PM_2.5_ and NO_2_ did not materially attenuate any of the four biomarker associations (IL-6, 1.18 vs. 1.20; D-dimer, 1.14 vs. 1.18; CRP, 1.14 vs. 1.14; fibrinogen, 1.14 vs. 1.13). There was no attenuation for CRP and fibrinogen, negligible attenuation for IL-6 (HR 1.20 to 1.18, an 11% reduction in excess risk), and modest attenuation for D-dimer (HR 1.18 to 1.14, a 22% reduction in excess risk). The attenuation of the D-dimer association suggests that air pollution may be a partial upstream determinant of hemostatic activation. However, the majority of the D-dimer–HF association (78%) persists after pollution adjustment, indicating that other structural exposures (e.g., neighborhood deprivation, occupational stress) also contribute to hemostatic burden. Short-term PM_2.5_ exposure is an established driver of cardiovascular hospitalization,^25^ and its inclusion here rules out ambient air pollution as a simple confounder of the biomarker–HF relationship. Rather, these results indicate that pollution operates as one upstream structural determinant among several—not the sole mediator of inflammatory immune tone.^25^ This pattern directs attention to the broader architecture of structural exposure, including neighborhood deprivation indices and psychosocial stressors, whose interaction with air pollution varies across neighborhoods and socioeconomic strata.^26^ Future analyses will operationalize these expanded structural exposure metrics to specify each pathway’s contribution to inflammatory immune tone and HF risk.

This study has several notable strengths. A pre-specified statistical standard operating procedure with prospective Holm family-wise error correction governed all confirmatory claims, eliminating the risk of post hoc inflation. The prospective multiethnic MESA cohort, with 16 years of follow-up and centrally adjudicated HF endpoints, is optimal for rare-outcome epidemiology. Linkage of geocoded air pollution (PM_2.5_ + NO_2_) with 91.3% high-accuracy geocode retention enabled explicit adjustment for environmental confounders, a capability unavailable in prior HF biomarker studies. Finally, multiple imputation (m = 20) with an imputation-versus-complete-case sensitivity analysis confirmed that the findings are not driven by differential biomarker missingness across racial groups.

Several limitations warrant consideration. Biomarkers were measured at a single time point (Exam 1), which may misclassify long-term inflammatory burden because of within-person variability and assay error, attenuating observed associations (regression dilution bias). Additionally, subclinical heart failure may have been present in some participants despite the exclusion of clinically manifest CVD. Reverse causation (subclinical HF elevating baseline inflammatory markers) cannot be fully excluded. At Exam 1, NT-proBNP was measured in 99.6% of participants, of whom 20.6% had elevated values (>125 pg/mL). Sensitivity analyses excluding participants with NT-proBNP >125 pg/mL or echocardiographic evidence of diastolic dysfunction are planned and will be reported in the supplementary materials. Additionally, immune tone is dynamic, and longitudinally patterned exposure may be more etiologically relevant than a single measurement. Fully adjusted models conditioned on BMI, hypertension, diabetes, and eGFR, which are plausibly downstream of chronic inflammatory burden, may induce collider bias and attenuate mediation estimates. Because these covariates may lie on the causal pathway between structural exposures, inflammatory burden, and HF, adjustment likely biases our estimates of mediation toward the null. Sensitivity analyses with minimally adjusted models (age, sex, race only) are planned to quantify the extent of this bias; preliminary estimates suggest that the mediation proportion may be higher in minimally adjusted models, consistent with collider-induced attenuation in fully adjusted models. The mediation analysis uses a sequential covariate-adjustment approach, which provides heuristic insight into how much of the Black–White hazard difference is statistically accounted for by biomarker adjustment but does not identify natural direct and indirect effects; formal counterfactual mediation is planned.

Additive interaction (RERI) analysis is ongoing, and the null multiplicative interaction result alone does not complete the equity science argument on the absolute risk scale. The air pollution sub-cohort (n = 6,476) is selected from geocodable participants; directional robustness was confirmed, but the sub-cohort may not fully represent the analytic sample. If geocodable participants differ systematically (e.g., by neighborhood stability or socioeconomic position), air pollution robustness analyses may under- or overestimate confounding; describing these differences in a supplementary table would aid interpretation. Finally, self-identified race/ethnicity is a social construct, not a biological variable; residual confounding by unmeasured structural determinants—including neighborhood deprivation, occupational exposure, and historical redlining indices—cannot be excluded with the current data linkage. Future linkage to neighborhood deprivation indices and redlining scores within MESA will be essential to quantify these residual pathways.

## Conclusion

In a prospective multi-ethnic cohort, four inflammatory and hemostatic biomarkers were independently associated with incident heart failure. A principal-components composite of inflammatory burden was a robust and biologically parsimonious predictor, and the inflammatory panel explained up to 57.5% of the inflammatory-mediated component of the Black–White HF risk gap. The absence of a multiplicative race × biomarker interaction indicates that per-unit biological risk is equitably shared across groups; the disparity resides in differential structural exposure. These findings, demonstrating (i) modest but consistent biomarker–HF associations, (ii) a composite index with an HR of 1.25 per SD, and (iii) up to 57.5% attenuation of the Black–White HF hazard difference in sequential models, reframe inflammatory biomarkers not merely as clinical risk predictors but as biological evidence of structural inequality: biomarker burden in minoritized populations is produced, not intrinsic. Prespecified next-phase analyses—formal causal mediation via CMAverse, RERI additive interaction on the absolute risk scale, and neighborhood-level structural exposure linkage—will complete the analytical framework. Equitable cardiovascular health requires structural determinants science: measuring biological burden is necessary but not sufficient without dismantling the structural conditions that produce it.

## Non-standard Abbreviations and Acronyms

BH: Benjamini–Hochberg
BMI: body mass index
CMAverse: R package for causal mediation analysis
CRP: C-reactive protein
eGFR: estimated glomerular filtration rate
FEU: fibrinogen-equivalent units
HF: heart failure
HR: hazard ratio
IL-6: interleukin-6
IQR: interquartile range
JAK-STAT3: Janus kinase–signal transducer and activator of transcription 3
LVEF: left ventricular ejection fraction
MESA: Multi-Ethnic Study of Atherosclerosis
NO_2_: nitrogen dioxide
NT-proBNP: N-terminal pro-B-type natriuretic peptide
PC1: first principal component
PCA: principal components analysis
PM_2.5_: fine particulate matter (aerodynamic diameter ≤2.5 μm)
RERI: relative excess risk due to interaction

## Acknowledgments

The authors thank the other investigators, the staff, and the participants of the MESA study for their valuable contributions. A full list of participating MESA investigators and institutions is available at http://www.mesa-nhlbi.org. This manuscript was prepared using MESA datasets obtained from the NHLBI Biologic Specimen and Data Repository Information Coordinating Center and does not necessarily reflect the opinions or views of the MESA or the NHLBI.

## Generative AI disclosure

Generative artificial intelligence was not used by the authors except to check for grammar and sentence-structure errors (Grammarly).

## Data Availability

The MESA data analyzed in this study are available to qualified investigators through the National Heart, Lung, and Blood Institute Biologic Specimen and Data Repository Information Coordinating Center (BioLINCC; https://biolincc.nhlbi.nih.gov) under a data use agreement. Codes to generate all tables and figures are available from the corresponding author via a public repository (https://github.com/riplrt/Mesa_Project).

